# Women’s experiences and acceptability of self-administered, home delivered, intravaginal 5-Fluorouracil cream for cervical precancer treatment in Kenya

**DOI:** 10.1101/2024.08.27.24312651

**Authors:** Konyin Adewumi, Aparna Ghosh Kachoria, Everlyn Adoyo, Mercy Rop, Antony Owaya, Jennifer H Tang, Lisa Rahangdale, Chemtai Mungo

## Abstract

**BACKGROUND:** Innovative strategies are essential to meet the World Health Organization’s 90/70/90 cervical cancer elimination targets, aiming for 90% access to precancer treatment globally by 2030. In low-and middle-income countries (LMICs) where most cervical cancer cases occur, access to precancer treatment is severely limited. Scalable solutions like self-administered topical therapies can help close this gap. In a recent Phase I trial (ClinicalTrials.gov NCT05362955), we demonstrated safety and adherence to self-administered intravaginal 5% 5-Fluorouracil (5FU) cream as an adjuvant therapy for cervical precancer among women living with HIV (WLWH) in rural Kenya. To understand women’s experiences with self-administered 5FU, we evaluated the acceptability of this intervention among trial participants.

**METHODS:** All 12 participants from the Phase I trial completed a structured questionnaire and in-depth semi-structured interviews in their preferred language, focusing on their experiences with 5FU self-administration, challenges faced, and overall acceptability of the intervention, including whether they would use it again or recommend it to someone who needed it. Quantitative data were analyzed using descriptive statistics. In the qualitative study, acceptability was defined as “the perception that a given treatment is agreeable, palatable, or satisfactory.” A thematic analysis was conducted using five dimensions of acceptability: content, complexity, comfort, delivery, and credibility.

**RESULTS:** The mean age was 43.9 years (SD 4.4), and seven (58%) had primary education or less. While some participants reported feelings of uncertainty when they started using 5FU, at the end of the study, all 12 participants strongly agreed that the cream was safe and were confident they used it correctly. Most participants (91.7%) experienced no discomfort with the vaginal applicator, and most reported using tampons overnight after 5FU use, as recommended. Qualitative findings revealed that favorable perceptions of self-administered 5FU were driven by its ease of use, the discrete nature of the treatment, and the comfort of home application. The main challenges included correctly measuring the study drug, finding a private place at home to self-administer, and the need to use condoms during treatment. Compared to their previous ablation or excision treatments, participants found 5FU to be less painful, and all would prefer a self-administered treatment instead of a procedure if it were an option.

**CONCLUSION:** Self-administered intravaginal 5FU as an adjuvant treatment for cervical precancer among women living with HIV in Kenya was highly acceptable. Randomized studies of 5FU and other topical therapies in LMICs are needed to evaluate their use in closing the current precancer treatment gaps in these settings.

## Introduction

Although cervical cancer is preventable, it is the second most common cancer among women worldwide.^1^ Global trends of cervical cancer represent a dire health inequity, with 85% of incident cases and 90% of deaths occurring in low- and middle-income countries (LMICs),^1^ due to lack of access to known primary and secondary prevention tools. The burden of cervical cancer is particularly pronounced in sub-Saharan Africa (SSA), a reflection of the lack of established health systems,^2^ and the dual epidemics of human immunodeficiency virus (HIV) and human papillomavirus (HPV).^3^ As a result, cervical cancer is the leading cause of preventable premature cancer deaths in LMICs.^4^ In 2020, the World Health Organization (WHO) launched the 90/70/90 global strategy to eliminate cervical cancer, calling for a (1) 90% HPV vaccination rate for girls, (2) 70% screening coverage, and (3) 90% of those diagnosed with cervical precancer or cancer adequately treated by 2030.^5^ Achieving these targets would help reach the WHO elimination threshold of four or fewer cases of cervical cancer per 100,000 women, averting 62 million deaths in LMICs alone.^6^ However, to achieve these targets, scalable strategies are needed to close the cervical precancer treatment gaps in LMICs.

Cervical cancer can be prevented through screening for and treatment of early changes in the cervix, also known as cervical precancer, cervical dysplasia, or cervical intraepithelial neoplasia grades I-III (CIN1-3), caused by HPV infection. Current cervical precancer treatment methods include ablation or excision procedures, both of which require trained healthcare professionals to perform.^7^ In LMICs, where the greatest burden of cervical cancer lies, access to precancer treatment is highly limited.^8–11^ In a 2021 review of Kenya’s national cervical cancer screening program, only 26% of 10,983 women with cervical precancer received treatment.^12^ Similarly, between 2011 and 2015 in Malawi, only 43% and 32% of women who required ablation or excision, respectively, received treatment.^11^ Challenges in treating cervical precancer in LMICs include; high costs, weak treatment infrastructure, a lack of skilled healthcare providers in rural facilities where most women live,^10,13^ privacy concerns associated with pelvic exams,^14–16^ and high loss-to-follow-up rates when women are referred to tertiary hospitals where treatment is available.^17^ A 2022 assessment of cervical cancer screening and treatment capacity in Kenya found that while 48% of 3,150 hospitals surveyed offered cervical cancer screening, only five percent offered treatment for precancerous lesions.^18^ Innovative, accessible, and resource-appropriate strategies are urgently needed to address these gaps in LMICs, including the use of self-administered intravaginal therapies that can be safely and effectively used at home.

While no topical therapies are currently approved for cervical precancer treatment, the use of self-administered topical treatments for HPV-associated anogenital lesions is an area of active investigation.^19–25^ The feasibility, safety, and efficacy of self-administered topical therapies for cervical precancer treatment have been demonstrated by several studies in high-income countries, including randomized trials.^23,26–28^ Some of the drugs currently being studied are on the WHO List of Essential Medications,^29^ a list comprised of medications considered necessary for all health systems and are usually accessible in generic form in LMICs. One such drug is 5-Fluorouracil (5FU) cream, which is an antimetabolite and cytotoxic drug used topically for treatment of various precancerous and cancerous skin conditions, including squamous cell carcinoma of the skin.^30^ In gynecology, topical 5FU cream has been used for treatment of genital warts^31,32^ and vulvar precancerous lesions.^33,34^ Two U.S randomized trials have demonstrated that self-administered intravaginal 5FU is a safe and effective treatment for cervical precancer.^26,27^ In a trial comparing intravaginal 5FU to six months of observation in HIV-negative women with cervical intraepithelial neoplasia grade 2 (CIN2), regression occurred in 93% of women treated with 5FU, compared to 56% in the observation group (p=0.01). ^27^ Both studies found that administering two grams of 5FU intravaginally every other week for eight applications was found to be safe, with primarily grade I adverse events (AEs), and no severe (grade III or higher) AEs.

Early studies in LMICs are exploring self-administered topical therapies for cervical precancer, including an ongoing trial in South Africa using 5FU as an adjuvant (additional) cervical precancer therapy among HIV-positive women.^35^ If proven feasible, acceptable, and effective, these therapies could provide a scalable and cost-effective treatment where provider-administered treatments are inaccessible. Topical therapies can be used as primary precancer treatment, as a bridge treatment when waiting times for an ablation or excisional procedure are long, or as an additional (adjuvant) treatment following ablation or excision to improve cure rates, especially for women living with HIV (WLWH) who experience high recurrence rates with current treatments.

Data on women’s experiences with and acceptability of topical therapies in LMICs are crucial for guiding efficacy and implementation studies of self-administered treatments for cervical precancer in these settings. However, there are no studies describing African women’s experiences with and acceptability of a self-administered intravaginal therapy for cervical precancer, following actual use. Recent qualitative^36^ and quantitative^37^ studies among Kenyan women undergoing cervical cancer screening or precancer treatment, and Kenyan men^38,39^ with female partners demonstrated high perceived acceptability of self-administered topical therapies. Among 376 women surveyed after cervical cancer screening, 191 (50%) with a history of cervical precancer treatment, 98% expressed willingness to use a self-administered intravaginal therapy for cervical precancer.^37^ In focus group discussions with 29 of these women, most of whom had undergone excision or ablation treatments, most participants favored a self-administered treatment, if it were available, citing increased access, less pain, and increased privacy compared to the treatment they received.^36^ However, in these studies, women had not actually used a self-administered therapy, hence their responses only reflected perceived acceptability. To address this gap, we used a quantitative and qualitative approach with the objective of exploring women’s lived experiences with and acceptability of self-administered intravaginal 5FU following a Phase I pilot clinical trial in Kisumu, Kenya.^24^

## Methods

### Clinical Trial Design and Setting

A Phase I pilot clinical trial was performed among women living with HIV (WLWH) in Kisumu County, Kenya, between March 2023 and February 2024 (ClinicalTrial.gov NCT05362955).^40^ Kisumu County is one of 47 administrative units in Kenya,^41^ a country of 55 million in East Africa.^42^ Kisumu County is among the highest HIV burden regions in Kenya, with an 18% prevalence rate, compared to a national average prevalence of 3.7 percent in 2022.^38, 43^ Cervical cancer is the second most common cancer among women in Kenya, contributing 5,250 (13%) of new cancer cases and 3,286 (12%) of all cancer deaths annually.^44^ Cervical precancer treatment is primarily ablation, with excision available in a few tertiary centers.

The trial, whose protocol is published,^40^ investigated repurposing 5FU cream as an adjuvant self-administered intravaginal therapy after cervical intraepithelial neoplasia grade 2/3 (CIN2/3) treatment in WLWH. 12WLWH, aged 18-49 years, with a history of CIN2/3, who were within four to 12 weeks after primary treatment with ablation or excision were enrolled. Participants had to be non-pregnant, use dual contraception (a long-acting method and condoms) if of childbearing age, and maintain sexual abstinence for 48 hours after each 5FU application. Details of the inclusion and exclusion criteria, participant recruitment process, and characteristics of study participants in this trial are provided in Supplemental Table 1, Supplemental Figure 1, and Supplemental Table 2. Participants self-administered two grams of 5FU intravaginally at home every other week for eight applications and were seen in the clinic on alternate weeks for safety and adherence assessments. Participants were advised to use the cream, which was dispensed via an applicator, at night before sleep, and to place a tampon vaginally following 5FU administration to keep the cream at the cervix. Participants were instructed to remove the tampon the morning after insertion, discard it, shower, and frequently wash their hands and change panty liners over the next three days to minimize irritation from the cream. Prior to treatment initiation, participants attended multiple education sessions with a study nurse. During these sessions, they learned how to load two grams of toothpaste onto a pre-marked applicator and used a pelvic model to practice advancing the applicator to the cervix and dispensing the cream. A competency checklist was used to confirm understanding of all self-administration steps before the study drug, provided in a 20-gram tube, was dispensed and participants self-administered the first dose in the study clinic. The primary outcome was safety, measured as the type, frequency, and severity of adverse events using a standardized grading scale.^45^ Secondary outcomes were adherence, tolerability, uptake, and acceptability. Adherence was evaluated using: 1) self-report, 2) weighing of study drug tube at each visit to document appropriate decreasing weight, and 3) visual inspection of returned applicators for evidence of exposure to the study drug. A participant was considered adherent to a specific 5FU dose if all three assessment methods supported its use. Tolerability was defined as the ability of participants to use all eight 5FU doses. Uptake was defined as the proportion of eligible women who agreed to participate in the trial. Acceptability was evaluated on week 16, using a questionnaire and in-depth interviews (IDIs). Safety and adherence results from the trial were previously published.^24^ The trial demonstrated that self-administered 5FU at the studied dose and frequency was safe, with primarily grade I adverse events (AEs) and no grade III or IV AEs. All 12 participants tolerated all eight 5FU doses, and adherence to 96% of the 5FU doses was supported by the three measures.

### Data Collection, Transcription, and Translation

Data on acceptability was collected from all 12 participants. Quantitative data was collected using a questionnaire adapted from a US study on self-administered 5FU.^27^ The questionnaire assessed participants’ motivation for joining the trial, their experience with study procedures, male partner support, privacy concerns with using the cream, and whether they would recommend the cream to others. The questionnaires were administered by trained research assistants in the participants’ preferred language, either English, Dholuo, or Swahili. IDIs were similarly conducted in local languages by qualitative research assistants from the community with prior experience conducting IDIs and familiarity with the local context. The IDIs were conducted using a semi-structured interview guide (Appendix A) specifically developed for this study. The discussions focused on participants’ experiences and confidence with self-administering 5FU, comfort with using an applicator as well as tampons, acceptability of the side effects, challenges to adherence, and male partner support, including the ability to maintain abstinence as required. The interviews were performed in a private room and were approximately 45 minutes. They were audio recorded, and the recordings were transcribed verbatim, translated to English, and crosschecked to confirm accuracy. Both the questionnaires and IDIs were conducted within Lumumba Sub-County Hospital in Kisumu, Kenya, where the trial took place.

### Sample Size

All 12 participants who took part in the Phase I trial were included in the survey questionnaire, as well as the in-depth interviews. The sample size considerations for the Phase I trial were previously described in the published study protocol.^40^

### Data Analysis

Quantitative data was entered directly into REDCap, a secure survey platform for analysis. Descriptive statistics were used to summarize questionnaire data. All statistical analyses were performed using R version 4.1.0 (Vienna, Austria). For qualitative data analysis, an initial codebook was created inductively, using the IDI guide to drive the creation of codes. This codebook was then amended by two coders (KA, EA) who read and coded three of the 12 transcripts to gain a sense of topics covered. Emerging themes were then added, deductively, to the codebook to create a final version. All IDI transcripts were then coded using the finalized codebook. To ensure inter-coder agreement, a subset of transcripts was randomly selected, and codes were compared for agreement; discrepancies were resolved through discussion and consensus, with revisions documented in the codebook. Analysis was performed using Dedoose version 9.2.012. Coding reports were generated from Dedoose and reviewed before analysis.

In this study, acceptability was defined as “the perception among a stakeholder or individual consumer that a given treatment, service, practice, or innovation is agreeable, palatable, or satisfactory.”^46^ This aligns with definitions in the literature, including in vaginal microbicide trials for HIV prevention, which include satisfaction with a product, willingness, and intent to use it, as well as actual correct and consistent use.^47–49^ According to Proctor et al., acceptability in implementation studies is assessed based on a consumer’s knowledge of or direct experience with various dimensions of a treatment or intervention and their satisfaction with aspects, such as its content, complexity, comfort, credibility, and delivery.^46^ To evaluate the acceptability of self-administered 5FU, we conducted thematic analysis and mapped the coded data onto these predefined dimensions, each with a specific definition that was agreed upon prior to the analysis (Table 1).

**Table 1.** Summary of qualitative findings by the dimension of acceptability.

| Dimension | Definition | Emerging Themes | Summary of Findings |
| --- | --- | --- | --- |
| Content | An assessment of women's perspectives on an intravaginal, self-administered topical therapy | Previous experiences with precancer treatment | As based on previous experience with precancer treatments, their educational background, and their reasoning for joining the study, women prefer the 5FU treatment better than LEEP, with specific instances of dislike for LEEP, indicating a preference for 5FU. |
| Complexity | An assessment of women's beliefs about how easy or difficult it was to use self-administered 5FU | Challenges with dispensing correct treatment dosage (amount). | Most women experienced initial trouble with the dosing, but with additional education, felt more comfortable with dosing, indicating minimal complexity with 5FU. |
|  |  | Treatment education as a facilitator to dispensing correct treatment dosage (amount). | All women said their confidence increased over time as they got more comfortable with the treatment. |
|  |  | Confidence using treatment | Some women credited the education they received in the clinic to their comfort and understanding. |
| Comfort | An assessment of women's experiences of and with adverse events, self-administering 5-FU, and using a tampon. | Adverse events | Most women mentioned experiencing some level of adverse events, but importantly, noted that this did not deter them from treatment since they knew to expect some level of discomfort initially. |
|  |  | The role of health care providers and research staff | Women who reported more severe adverse events appreciated being able to talk to a nurse, who would help them through it or encourage them to stay on treatment. |
| Credibility | Women's beliefs around the efficacy of the treatment | Hope and faith | Women believed that the treatment would work and expressed a lot of hope in the treatment as a solution. |
| Delivery | Assessment of women's perspectives on aspects of treatment | Preferred location of treatment delivery | Most women preferred home delivery for privacy, ease of life (not interfering with work/ not needing to go to clinic). |
|  | delivery; where (home or clinic), when (time of day), and other barriers to use | Timing of self-administered treatment | Most women the ability to use the treatment more discretely at night w/ discussing their preference for home delivery. |
|  |  | Barriers to self-administration | Some women did indicate lack of privacy concerns when their kids were out of school. |

### Ethical Considerations

The study was approved by the ethics review boards at Maseno University School of Medicine in Kenya (MUSERC/01136/22) and the University of North Carolina at Chapel Hill (IRB number 22-1978) in the United States. All participants provided consent prior to study participation.

## Results

### Participant Characteristics

The mean age was 43.9 years (SD 4.4). Seven (58%) participants had a primary education or less as the highest level of education attained. Only three (25%) were formally employed, while the rest were peasant farmers (n=4, 33%), self-employed (n=2, 17%), or unemployed (n=3, 25%). Five (42%) participants were married. All 12 had a monthly income of less than Kenya Shillings 25,000 (∼ US dollars 120), were on antiretroviral therapy, and had prior treatment with ablation or excision for high-grade cervical precancer.

### Quantitative findings

All 12 participants completed the acceptability questionnaire (Table 2). While some participants reported feelings of uncertainty when they started using 5FU, including worry and anxiety (n=3, 25%), or fear (n=2, 17%), at the end of the study, all 12 strongly agreed that the cream was safe and were confident that they used it correctly. When asked about the most difficult part of using the drug, four participants (33%) noted negotiating partner support, two (17%) mentioned the side effects and the need to use condoms for contraception, and one (8%) noted the need to abstain from sex after 5FU use. However, all six participants who had a sexual partner during the study strongly agreed that their partner ultimately supported their use of 5FU cream. Most participants, 11 (91%), noted no discomfort with using the applicator to insert the study drug, while one (8%) participant noted mild discomfort with the vaginal applicator. The use of tampons overnight after applying the cream was common, with 11 participants (92%) reporting they used tampons each time they used 5FU. All 12 participants strongly agreed that they were comfortable using tampons; however, five (42%) strongly agreed that using a tampon made it harder to use the study drug. All 12 participants reported that they would prefer to use the cream for primary precancer treatment if given the option, compared to undergoing the ablation or excision procedure they had.

**Table 2:** Acceptability of self-administered intravaginal 5-Flourouracil cream for cervical precancer treatment among HIV-positive women in Kenya (n = 12)

|  |  |
| --- | --- |
| **1. Motivation for participating in the study<sup>1</sup>** | **n (%)** |
| To contribute important information to medical science | 8 (66.7%) |
| To help other patients with the same condition | 9 (75.0%) |
| To receive treatment for this condition | 11 (91.7%) |
| To benefit from the additional medical attention and testing that the study provided | 12 (100.0%) |
| To benefit from the financial reimbursement offered | 5 (41.7%) |
| **2. Emotions or feelings when you first used the cream<sup>1</sup>** |  |
| Anxious or worried | 3 (25.0%) |
| Embarrassed or shame | 0 (0.0%) |
| Afraid or fearful | 2 (16.7%) |
| Relieved | 0 (0.0%) |
| Empowered or confident | 4 (33.3%) |
| Overwhelmed | 0 (0.0%) |
| Did not feel anything at all | 3 (25.0%) |
| Other | 0 (0.0%) |
| Don't know, no opinion | 1 (8.3%) |
| **3. Most difficult part about using the cream<sup>1</sup>** |  |
| None | 8 (66.7%) |
| Finding time to apply it | 0 (0.0%) |
| Finding privacy to apply it | 0 (0.0%) |
| Inserting the cream with the applicator | 0 (0.0%) |
| Having to place a tampon overnight following inserting the cream | 0 (0.0%) |
| Being sure that I put the cream in correctly | 0 (0.0%) |
| Remembering to put in the cream | 0 (0.0%) |
| Having to abstain from sex for two days after using the cream | 1 (8.3%) |
| Side effects of the medication | 2 (16.7%) |
| Getting my partner's support to use the cream | 4 (33.3%) |
| The need to use condoms for contraception | 2 (16.7%) |
| **4. Physical discomfort with use of the applicator** |  |
| None | 11 (91.7%) |
| Mild | 1 (8.3%) |
| Significant | 0 (0.0%) |
| Not sure | 0 (0.0%) |

<sup>1</sup> Multiple responses allowed

|  |  |
| --- | --- |
| **5. Frequency of tampon use after applying the cream** |  |
| Always (each time) | 11 (91.7%) |
| Most of the time | 1 (8.3%) |
| Fewer than half the time recommended | 0 (0.0%) |
| Never | 0 (0.0%) |
| **6. I was comfortable using the tampon overnight after drug application** |  |
| Strongly agree | 12 (100.0%) |
| Somewhat agree | 0 (0.0%) |
| Don't know, no opinion | 0 (0.0%) |
| Somewhat disagree | 0 (0.0%) |
| Strongly disagree | 0 (0.0%) |
| **7. The need to use a tampon to keep the medication in place overnight made it harder for you to use the cream?** |  |
| Strongly agree | 5 (41.7%) |
| Somewhat agree | 0 (0.0%) |
| Don't know, no opinion | 0 (0.0%) |
| Somewhat disagree | 0 (0.0%) |
| Strongly disagree | 7 (58.3%) |
| **8. Prefer self-application at home to clinic application by a doctor** |  |
| Would strongly prefer self-application | 11 (91.7%) |
| Would likely prefer self-application | 1 (8.3%) |
| Would likely prefer the doctor to apply it | 0 (0.0%) |
| Would strongly prefer the doctor to apply it | 0 (0.0%) |
| Not sure | 0 (0.0%) |
| **9. You were discouraged from using this vaginal cream by friends and family** |  |
| Yes | 1 (8.3%) |
| No | 11 (91.7%) |
| Don't know, no opinion | 0 (0.0%) |
| **10. It was hard to remember to use the cream** |  |
| Strongly agree | 0 (0.0%) |
| Somewhat agree | 0 (0.0%) |
| Don't know, no opinion | 0 (0.0%) |
| Somewhat disagree | 0 (0.0%) |
| Strongly disagree | 12 (100.0%) |
| **11. It was hard to find privacy to use the cream** |  |
| Strongly agree | 0 (0.0%) |
| Somewhat agree | 0 (0.0%) |
| Don't know, no opinion | 0 (0.0%) |
| Somewhat disagree | 1 (8.3%) |
| Strongly disagree | 11 (91.7%) |
| **12. Thinks the vaginal cream is safe** |  |
| Strongly agree | 12 (100.0%) |
| Somewhat agree | 0 (0.0%) |
| Don't know, no opinion | 0 (0.0%) |
| Somewhat disagree | 0 (0.0%) |
| Strongly disagree | 0 (0.0%) |
| **13. Confident that I used the vaginal cream correctly** |  |
| Strongly agree | 12 (100.0%) |
| Somewhat agree | 0 (0.0%) |
| Don't know, no opinion | 0 (0.0%) |
| Somewhat disagree | 0 (0.0%) |
| Strongly disagree | 0 (0.0%) |
| **14. Partner was supportive of my using the vaginal cream** |  |
| Strongly agree | 6 (50.0%) |
| Somewhat agree | 0 (0.0%) |
| Don't know, no opinion | 0 (0.0%) |
| N/A (Does not have a partner) | 6 (50.0%) |
| **15. Would recommend 5FU use to others** |  |
| Strongly agree | 12 (100.0%) |
| Somewhat agree | 0 (0.0%) |
| Don't know, no opinion | 0 (0.0%) |
| Somewhat disagree | 0 (0.0%) |
| Strongly disagree | 0 (0.0%) |
| **16. In the future, if you need this cream as part of your treatment, would you use it again?** |  |
| Strongly agree | 12 (100.0%) |
| Somewhat agree | 0 (0.0%) |
| Don't know, no opinion | 0 (0.0%) |
| Somewhat disagree | 0 (0.0%) |
| Strongly disagree | 0 (0.0%) |
| **17. If the cream could be used alone to treat precancer, would you prefer using the cream and applying it at home as you did, or would you prefer the treatment procedure you had?** |  |
| Would strongly prefer the cream instead of the procedure | 12 (100.0%) |
| Would likely prefer the cream instead of the procedure | 0 (0.0%) |
| Would likely prefer the procedure instead of the cream | 0 (0.0%) |
| Would strongly prefer the procedure over the cream | 0 (0.0%) |
| Don't know, no opinion | 0 (0.0%) |

### Qualitative findings

Findings from the qualitative analysis are summarized by each dimension of acceptability, whose definitions are shown in Table 1 and discussed below.

### Content

To assess the acceptability of the content of the intervention, we assessed women’s perspectives of an intravaginal, self-administered topical therapy for cervical precancer treatment. Participants compared their experience with self-administered 5FU to their prior experiences with ablative or excisional precancer treatment, which they had all undergone. During the interviews, most participants expressed a preference for 5FU over the previous treatments, including loop electrosurgical excision procedure (LEEP).

> “When I was done for LEEP procedure after that I could not do my work, but for this treatment [5FU], [I] am able to do my work as usual as I was using the 5FU treatment, but when l was done [with] LEEP, I could not do my work… 5FU treatment is good because it does not interfere with your daily work.” - R12

Some participants reported less fear with a self-administered treatment compared to conventional provider-administered treatments:

> “I found that LEEP to be something that people can fear but this treatment (5FU) is something you just insert and relax” – R05

> “It’s easy to use [5FU cream] because when I compare it with LEEP someone can be scared and run away without treatment (laughing)…No, LEEP is a long journey…”- R06

Participants’ confidence with and preference for 5FU self-administration also stemmed from the education that they received from the study staff, who felt they effectively informed them about 5FU use.

> “I was happy with this treatment because when I went for treatment, the doctors were talking to us well and they were teaching us how to continue using the cream.”- R04

> “Following the information I was given, how I was going to use the treatment, this encouraged me, I went to the clinic before I was given enough information, when I reached there then I was given detailed information, that’s why I decided to participate in the study because they taught me how I was going to use the cream and other information about the cream and this brought home to me about the treatment.” - R10

Several participants highlighted the frequency of 5FU use as something they liked, which was once every two weeks, which was felt to be easy and not burdensome:

> “It was good because it’s something you insert and then you are done, it’s not something you would say it was used daily that you can forget, it’s something you use once after two weeks, so it was easy to use” - R06

Participants also noted certain aspects of using 5FU that they disliked, including the need to use condoms for dual contraception to prevent pregnancy, which in some cases resulted in disagreements with spouses, as well as some of the adverse events they experienced, such as vaginal itching, burning, and discomfort during urination:

> “When am being screened [pelvic exam], so this is what I didn’t like and also the issue of using condoms brought an issue between me and my husband, so its these two that I didn’t like.”- R04

> “The treatment was good, but it had side effects because after using the cream maybe after three to four days l could feel the place was itching and spilling out, so I used to take warm salty water and then I clean with then I sprinkle water after urinating because it was paining after urinating.” - R04

### Complexity

To evaluate the complexity of the intervention, we assessed women’s beliefs about how easy or difficult it was to use self-administered 5FU. As part of 5FU self-administration, participants were instructed to attach the 40-gram tube of 5FU cream to a provided pre-marked applicator, dispense two grams of 5FU into the applicator, insert the applicator intravaginally until they meet resistance, and dispense the medication. Emerging themes included difficulties in measuring the correct dosage (two grams), and the impact of the education they received on increasing their confidence in their ability to self-administer correctly.

Several participants expressed challenges with using the applicator and being confident that they measured the drug correctly.

> “But [the] applicator was difficult to use if you were not understanding, for the first time I could not use it well, yes because, so here am requesting [the] researcher that if its 2 grams, there should be a barrier to that level which when I pull the cream then I don’t surpass.” - R02

> “It was a little bit of a challenge because measuring the cream [so] it was not to pass some level, that’s where I had a challenge [with], but I later learned.” - R10

Despite initial challenges, participants noted that their confidence in self-administration increased over time, often crediting the education they received from the study staff.

> “Using the cream at first was a problem but as I continued using it, I gained confidence, when I started, I was measuring with fear, so when I came and told that I didn’t measure the cream in the correct way and when I went back, I gained confidence of using the cream and used it well.” - R07

Other aspects of complexity with 5FU self-administration included limited knowledge of female anatomy among some participants such as being unsure about their ability to access the cervix:

> “Using it was challenging because during my growth [I] have never known if there is two parts in the vagina, one for urinating and another for birth, so when I was asked to insert this cream and I was wondering how I was going to apply this cream and urinate at the same time, so when I asked then I was told that there are two places, one for urinating and another one for birth. Where I had a challenge was when I started, the challenge was identifying where the cervix was, I had no one to ask but just to fix it on my own, that was the challenge I had but after using the cream for a while then it was not a challenge anymore.” - R06

Some participants had challenges using a tampon for the first time, including concerns about whether it would stay in place after insertion. However, regular visits to the study clinic, where instructions were reinforced and questions answered, also increased their confidence around tampon use:

> “Using the cream at home, when I first started, I felt it was difficult because I felt like how will I apply this cream to be inside, I thought I might insert the cream and when I go for short call [urination] then the tampon can come out and this was at first, and as I continue using it, because when I go to the hospital I used to ask things that were not clear to me and later I got it was an easy process even when I go for short call, the tampon remained inside, so I didn’t feel any challenge using the treatment at home.” -R12

However, not all participants expressed difficulty using tampons.

> “I had no problem using a tampon because I was told that when I use tampon then it will block the cream from coming out, so I see it was helping the cream to be inside.”- R02

While tampons were provided in the study, a participant noted that if women had to purchase them, it would be difficult for many, highlighting a potential barrier to treatment:

> “Just that tampon… for me I can buy but other people might not be able to buy, when talking I talk when having other people in mind too, because people are not the same.” - R08

### Comfort

To evaluate comfort, we assessed women’s experiences with adverse events associated with the treatment and their experience of using tampons as part of the process. Many participants expressed a strong preference for the 5FU cream over the previous precancer treatment they received. Reasons include less pain compared to the LEEP procedure which many had undergone:

> “If I compare this treatment [5FU cream] with the one, I was done for first. LEEP was very painful, l felt a lot of pain, but this one was good because the more I continued using it, I was getting used to it and to get to know how to use it, and using it was now easy. The little problem l had was not that big that could hinder me from doing my work, so using the cream was good to me compared to the first treatment I was done for called LEEP.” – R07

> “After I was done for LEEP, I felt pain for a long time, about three months when I could not do heavy duties, even walking for long was not easy…I would recommend [other women] to use 5FU cream because it works without pain, after using the cream as instructed by the doctor, I didn’t experience anything bad.” - R10

Most participants experienced some adverse events, including vaginal discharge, itching or discomfort with urination around the time of 5FU use. When asked about the impact of the side effects, a participant noted that the education she received during counseling provided her with an adequate understanding of the side effects and how to manage them, which increased her confidence in using 5FU:

> “When I was starting the treatment, I was told that I might experience the side effects, so it’s, something I knew that might happen, but they are things that might not harm someone and incase its severe you call the doctor, they tell you the way forward, so I wasn’t scared.” - R06

Another participant normalized the side effects she experienced, which included increased vaginal discharge, comparing them to expected side effects of malaria treatment which she had experienced:

> “Yes, I just like the treatment because even for example chloroquine when you use it sometimes you can feel your body itching, so you can’t stop using it, the key thing is to be healed. So those were just side effects, but treatment is ongoing.”- R07

Many participants emphasized that the education and teaching provided by the study nurses and clinic team significantly increased their comfort with using the treatment.

### Credibility

To evaluate credibility, we assessed women’s beliefs around the efficacy of 5FU as a precancer treatment. In counseling, participants were told that 5FU was being studied to be used as an additional therapy to improve precancer outcomes in WLWH following ablation or excision, and it may have a role as a primary treatment for some women.

When assessing the credibility of the treatment, hope and faith emerged as prevalent themes. Many participants believed that the treatment would be effective, and they felt hopeful, especially due to their regular interactions with the study clinical team during safety visits.

> “I was happy with this treatment because when I went for treatment, the doctors were talking to us well and they were teaching us how to continue using the cream.”- R02

> “What I liked about it was the visits to the clinic after using the treatment because they were checking if I have any problem and the doctor would advise.” - R04

Another participant noted that her confidence increased from the education she received during the study:

> “I gained some knowledge that I can share with others about the treatment and its goodness”- R12

Another participant expressed hope that using 5FU would help prevent a recurrence of precancer and prevent her from getting her uterus removed, which was recommended in the case of a recurrence:

> “When I was called to join the study, I felt good and hopeful…That I was going to get treatment and when I go back for test, I will be negative, that’s why I accepted easily because I felt that the hysterectomy procedure that was to be done will now not be done and I was happy for this… and I said that if this drug can help me then its better, so I want to have faith that this cream will help me.”- R10

### Delivery

To evaluate acceptability of the delivery dimension of this intervention, we assessed women’s perspectives on the location of 5FU use (home or clinic), timing of use, and other barriers to use. A key factor in the acceptability of the 5FU treatment, as highlighted in the interviews, is its ease of delivery – specifically, the ability to self-administer at home. Participants repeatedly commented on this as a reason they liked the treatment:

> “Using the cream at home, the reason why l liked using it at home is because after using it at home, I was going nowhere at night I could just sleep and the cream just work.”- R04

> “Because [at home] I was very free and alone, compared to the hospital which is open and many people are available, after am done with all my work [at home], then I use it when am alone in my bedroom.” - R10

A participant noted that a treatment used at home prevents disruptions in her daily activities that would occur if she needed to come to the hospital for treatment:

> “Because when using it at home as I said earlier it gives me time to do other activities during the day, because like when am here in the hospital the activities that I was to do during the day has stopped but when am at home I continue with my other duties.” -R06

Self-administration at home also enabled use at night, which many women preferred, and felt was easier to remember:

> “Using medication at home, I liked it because I was using the medication at night after doing all my work, when I apply the cream at night that’s my sleeping time so that’s why l liked it because I was using it at night, if I were to use it during the day, I might have forgot sometimes, after doing my daily activities, so at night was okay.”- R09

> “When I heard it was to be done at night then I was comfortable, because during the day sometimes you have a customer and women are always busy.” - R02

Others highlighted increased privacy with self-administration at home, which does not require a pelvic exam by a provider:

> “When using it in the hospital the doctors and nurses has to be there, and I would not like them to see my private part…during the time they examine me.” - R09

However, some participants noted that having privacy at home was a challenge they had to overcome:

> “Using the cream at home, sometimes when you want to apply the cream and my husband is in the house, I would not be free to use it.” - R04

> “Sometimes [the] time to use the cream has reached and you now have to tell people to go out and leave space for you to use the cream, sometimes you want to use the cream, and people are still in the room talking, so you feel like you don’t have that freedom.” - R05

Privacy was an issue for participants who lived in a one-room households, especially when their children were around. They noted difficulty with finding a private place to self-administer the drug:

> “When kids closed school, the room that I was using was full, using the cream was now a challenge, I had to use the cream inside the toilet for the children not to see me when using it, sometimes I had to wait until the kids go out to enable me use the cream when they are not in the room.” - R09

## Discussion

In this study on women’s experiences and acceptability of self-administered intravaginal 5FU following a feasibility clinical trial in Kenya, we found that participants who had previously undergone ablation or excision for high-grade precancer highly supported the use of self-administered 5FU as a precancer treatment. In quantitative findings, some participants, more than half of whom had a primary education or less, reported initial feelings of anxiety or fear when first using the cream. However, after completing a 16-week course of 5FU self-applied once every two weeks, all 12 participants were confident they used the cream correctly. All participants stated they would use 5FU again for precancer treatment if recommended and would also recommend it to other women needing such treatment. Importantly, all participants expressed a preference for self-administered 5FU over the ablative or excisional treatments they had received, if it were available as a primary precancer treatment. Qualitative findings largely supported the quantitative data, with participants specifically highlighting the ease, reduced pain, and decreased fear associated with a self-administered treatment compared to the procedures they had undergone. They also valued the privacy and convenience of a home-administered therapy, which caused less disruption to their daily lives. However, some participants had challenges with certain self-administration procedures, which required multiple education sessions to gain confidence.

To our knowledge, this is the first study to describe African women’s experiences and acceptability of self-administered 5FU for cervical precancer treatment. These findings of high acceptability are consistent with the pilot trial results, which demonstrated excellent adherence to self-administered 5FU at the dose and frequency studied,^24^ as adherence has been described as a component of acceptability.^47,50^ Our findings, though based on a small pilot trial, support continued feasibility studies on the use of topical therapies for cervical precancer treatment in LMICs and underscore the potential of these therapies to address the significant treatment gaps in these settings. If supported by randomized efficacy trials, self-administered topical therapies could have several uses in LMICs, where 90% of cervical cancer cases occur and provider-administered therapies are often inaccessible. They could serve as primary precancer treatment, as has been demonstrated in several randomized trials.^23,27^ They can be used as an interim treatment while awaiting definitive treatment at regional centers, where procedures like excisions are performed. This approach could be particularly impactful given the significant delays between referral from rural areas and successful treatment at tertiary centers – a median wait time of 167 days (5.5 months) in a study from Kenya.^51^ During this period, women are at risk of lesion progression or being lost to follow-up. Additionally, studies in these settings demonstrate that 32% - 60% of women with precancerous lesions referred to treatment do not attend their appointments.^8,12,13^ Starting a topical therapy immediately following diagnosis could be crucial as programs work to strengthen referral pathways. Lastly, topical therapies can be used as adjuvant therapy following primary treatment for those at high risk of recurrence, including WLWH or women with positive margins after excision.^52^

In our trial, participants were WLWH who had biopsy-confirmed CIN2/3 and had undergone primary treatment with ablation or excision and have been shown to have a 19-30% CIN2+ recurrence rate at 24 months.^53^ A U.S-based randomized trial demonstrated that WLWH randomized to self-administered 5FU following primary CIN2/3 treatment had eight percent recurrence at 18 months, compared to 31% in the observation arm (p=0.014).^26^ This demonstrates the potential of self-administered 5FU as an accessible strategy to improve precancer treatment outcomes in WLWH in SSA who are at the highest risk of cervical cancer.

Several insights from our findings can inform future studies on women’s use of self-administered therapies in LMICs. In qualitative interviews, many participants expressed hesitancy about their ability to correctly measure the appropriate amount of the study drug into the pre-marked applicator despite initial education sessions. Practical challenges encountered during the trial included inadequate lighting at home at night, making it difficult for some to clearly see the two-gram mark, and a participant with poor eyesight who required assistance. However, after repeated education sessions with a study nurse, most women gained confidence with this procedure. To address challenges with accurate measurement, future studies and implementation trials can use applicators pre-filled with the correct amount of study drug, although this may increase costs. The need for adequate patient education to ensure the safe and correct use of topical therapy was also highlighted by several participants who described a limited understanding of female anatomy. In the study, the use of pelvic models was critical for education and counseling, including on how to access the cervix. In larger trials, participant education can be enhanced through pre-recorded videos, regular text messages answering frequently asked questions, and the use of peer educators, which has been explored to improve uptake and adherence in other fields.^54^ Another important finding is that while all participants favored home use of 5FU, some faced challenges in finding a private place in the home to self-administer, including those living in single-room homes. Future trials can explore the impact of this on uptake and adherence and consider strategies to mitigate it, including whether to discuss use with others in the home.

Women’s preferences for and acceptability of intravaginal interventions have primarily been studied in the fields of contraception and HIV prevention.^55,56^ Within the vaginal microbicides for HIV prevention literature, product acceptability has been shown to be multifactorial, depending on packaging, side effects, safety, ease of use, and impact of sexual pleasure, among other factors.^55,57^ As was shown in the field of vaginal microbicides, product acceptability is a crucial driver of adherence,^55^ which impacts efficacy. Unlike in contraception or HIV prevention, topical therapies for precancer treatment would be used in the setting of treatment and not prevention. Adherence will be crucial for treatment efficacy in this context to prevent progress to cancer, making it essential to understand the drivers of adherence, including the impact of acceptability on adherence.

While our study is the first to describe African women’s experiences and acceptability of self-administered 5FU for cervical precancer treatment, a few studies have evaluated the acceptability of topical 5FU for cervical precancer in high-income settings. In a US randomized trial of topical 5FU for primary treatment of CIN2, acceptability, as reflected by adherence to the regimen, was high – 86% of participants reported using all eight doses, with a self-reported mean of 7.5 out of eight applications.^27^ In this study, while minor side effects were common, 83% reported feeling satisfied with the use of 5FU cream, 97% believed the cream was safe, and all participants in the 5FU arm were confident they had used it correctly. Despite significant differences in geography (US vs. Kenya), median age (24 years vs. 44.5 years), and education levels (74% vs. eight percent with more than a high-school education) between the two studies, the acceptability of 5FU was similarly high.

Most geographically comparable to our study is a 2016 single-arm, proof-of-concept trial of a self-administered antiviral vaginal pessary for cervical precancer treatment among HIV-negative women in Kenya.^58^ In this trial with 23 participants, acceptability, defined as adherence, was described as good, as no participants skipped a dose due to side effects or because their partner wanted intercourse during the two weeks of daily use of the study drug. This finding is important, as requirements to abstain from sex during the use of topical therapies can impact safety, adherence, and acceptability, particularly in situations where women are not able to negotiate sex or condom use.^42^ Because of the pro-inflammatory nature of potential intravaginal therapies for cervical precancer, abstinence for specific periods around their use is recommended.^26,27,34^ Inability to adhere to abstinence recommendations while using these therapies may be associated with worse side effects, potentially increasing a woman’s risk of contracting sexually transmitted infections, including HIV,^27^ and may expose the partner to the agents in case of barrierless intercourse.

In our trial, condom use was required during the duration of the trial for dual contraception because 5FU is a teratogen. Both qualitative and quantitative findings highlighted that this requirement was challenging, particularly for married couples. Future studies should focus on strategies to effectively communicate the importance of condom use to male partners to improve adherence and uptake. In this trial, 5FU was used once every two weeks, with abstinence required only for 48 hours after application. As a result, maintaining abstinence was not commonly reported as a barrier to use and acceptability among partnered participants.

However, topical therapies requiring daily application and sexual abstinence may face lower acceptability among women who cannot negotiate sex, including sex workers who are at increased risk of HPV.^59^ Future larger studies with greater statistical power can investigate whether women can adequately adhere to recommendations of condom use and abstinence and any adverse events associated with the inability to do this. Of note, there were no pregnancies during the study time frame, indicating that women can prevent pregnancy if needed for treatment.

Our study has several limitations, including the small sample size, being a pilot and single-site study. Future larger trials with multiple sites can provide more evidence of the acceptability of topical 5FU for cervical precancer in different contexts. The strengths of our study include reporting acceptability following the actual use of 5FU, which is a more accurate depiction than studies evaluating theoretical acceptability. Secondly, our study population of WLWH is a potential target population for topical therapies; hence, their inclusion is timely.

In summary, our pilot trial demonstrates high acceptability of self-administered intravaginal 5FU as an adjuvant treatment for CIN2/3 among WLWH, with participants preferring it over ablation or excision if recommended. Our findings underscore the need for ongoing research into topical, self-administered therapies for precancer treatment in LMICs, where the burden of cervical cancer is greatest. A randomized study is currently underway in South Africa among WLWH to evaluate whether self-administered intravaginal 5FU following excision for CIN2/3 will reduce recurrence rates (5R01CA250850). Similar studies are planned in LMICs to assess use of self-administered 5FU as primary precancer treatment in LMICs among both WLWH and HIV-negative women, aiming to expand access to precancer treatment. If randomized studies confirm efficacy of self-administered 5FU in these settings, its affordability and availability as a generic medication could enable distribution through community pharmacies that are accessible to women, including in rural areas. Self-administered precancer treatment could address barriers such as limited human resources, strained health systems, and geographical challenges in these settings, potentially saving millions of lives and contributing to the elimination of this preventable cancer.

## Data Availability

Anonymized research data are available upon reasonable request.

