## Appendix A_5FU_IDI English_Guide_Final for "Women’s experiences and acceptability of self-administered, home delivered, intravaginal 5-Fluorouracil cream for cervical precancer treatment in Kenya": Appendix A_5FU_IDI English_Guide_Final.docx (from another version of this preprint: version 1, 2024-08-28)

### In-depth Interview Guide

#### Introduction

*Good morning/afternoon, my name is …….,*

*Thank you for participating in this study. I’m going to ask you some questions about your personal experiences with the 5FU treatment. You are the expert of your own experiences, so there are no right or wrong answers. If you don’t feel comfortable answering any of the questions, please let me know, and we can either skip the question or come back to it later. My main goal is to learn from you and to have you speak freely. Remember that your answers will not be associated with your name, so you can feel comfortable and be as honest as possible. We will record your responses to help us with analysis, but your name will not be included in the answers so it cannot be linked to you in any way.*

*Do you have any questions before we begin?*

*I am now going to begin recording. You may ask me to stop recording at any time.*

#### **START the recording**

#### Interview Questions

##### Background

1. Before this study, had you used any vaginal medication at home?
   - If yes, what was the treatment?
2. What went through your mind when you were invited to participate in this study?
3. Why did you decide to participate?

##### Experiences using 5FU

*Now I’d like to ask you some questions about your experiences using the 5FU treatment.*

4. What did you think of using this treatment?
   - What did you like about it?
   - What did you not like?
   - Is there anything you wish could have been different?
   - Any suggestions for improving it?
5. How did you feel about self-administering the treatment at home?
   - Were there any specific challenges you faced? (e.g., privacy, understanding how to use the medication…)
   - What did like about it?
   - What did you not like?
6. How confident did you feel using this treatment?
   - Did that change over time? Tell me more about that.
7. Did you experience any side effects from the treatment?
   - Can you tell me about them?
   - Probe on severity, acceptability..
8. How was it using a tampon for this treatment?
   - How often did you use it?
   - What went well using it?
   - Any challenges?
9. How did your partner respond to your use of this treatment?
   - Did he have any concerns?
   - What was his response to needing to abstain from sex?
10. Are there other challenges you faced in using this medication at home that we have not discussed? If so, please discuss them.

*As you know, we are studying how this medication can be used to improve HPV or cervical precancer treatment for women in Kenya and Africa. Now I’d like to get your thoughts on using 5FU if it were to become available outside of a research study.*

##### Future Use

11. Based on your experience, if this medication becomes available for women with cervical pre-cancer to use, would you consider using it again?
    - Why or why not?
12. If you were to use this medication in the future, would you prefer to use it at home or have it administered by a healthcare professional at a clinic?
    - Why?
13. Would you recommend this medication to other women in a similar situation?
    - Why or why not?

##### Closing

14. Any other questions or thoughts you have?

*Thank you for talking with me today.*
